# Development of a clinical risk score to risk stratify for a serious cause of vertigo: A prospective cohort study

**DOI:** 10.1101/2024.03.04.24303562

**Authors:** Robert Ohle, David W. Savage, Danielle Roy, Sarah McIsaac, Ravinder Singh, Daniel Lelli, Darren Tse, Peter Johns, Krishan Yadav, Jeffrey J. Perry

## Abstract

**Objectives:** Identify high-risk clinical characteristics for a serious cause of vertigo in patients presenting to the emergency department.

**Design:** Multicentre prospective cohort study over 3 years.

**Setting:** Three university-affiliated tertiary care emergency departments.

**Participants:** Patients presenting with vertigo, dizziness or imbalance. A total of 2078 of 2618 potentially eligible patients (79.4%) were enrolled (mean age 77.1 years; 59% women).

**Main outcome measurements:** An adjudicated serious diagnosis defined as stroke, transient ischemic attack, vertebral artery dissection or brain tumour.

**Results:** Serious events occurred in 111 (5.3%) patients. We used logistic regression to create a 7-item prediction model: male, age over 65, hypertension, diabetes, motor/sensory deficits, cerebellar signs/symptoms and benign paroxysmal positional vertigo diagnosis (C-statistic 0.96, 95% confidence interval [CI] 0.92–0.98). The risk of a serious diagnosis ranged from 0% for a score of <5, 2.1% for a score of 5-8, and 41% for a score >8. Sensitivity for a serious diagnosis was 100% (95% CI, 97.1-100%) and specificity 72.1% (95% CI, 70.1-74%) for a score <5.

**Conclusions:** The Sudbury Vertigo Risk Score identifies the risk of a serious diagnosis as a cause of a patient’s vertigo and can assist physicians in guiding further investigation, consultation and treatment decisions, improving resource utilization and reducing missed diagnoses.

## Introduction

Vertigo is a common and costly reason for emergency department visits. Studies have shown that patients use the terms dizziness, vertigo and imbalance interchangeably to describe their symptoms of dizziness.^1^ We use the term vertigo to encompass vertigo, imbalance and dizziness.

It is the third most common reason for emergency department visits, resulting in significant resource utilization.^2–4^ Of these patients, only 2-5% will have a serious cause for their vertigo. The most common serious causes of vertigo include stroke, transient ischemic attack (TIA), brain tumour and vertebral artery dissection.

Patients presenting with vertigo have a higher rate of investigation and emergency department length-of-stay than non-vertigo patients.^5,6^ A large proportion of vertigo patients (30-50%) undergo a computed tomographic (CT) scan of the head, 98% of which are negative.^7^ Head CT is limited in investigating patients with acute stroke and TIA, given its extremely low sensitivity (7%–16%).^8^ Despite a high investigation rate, a population cohort study found that patients discharged with a benign dizziness diagnosis had a 50-fold increased risk of being admitted to the hospital within seven days with a stroke diagnosis compared to matched controls.^9^

Physicians lack validated clinical guidelines to help them make diagnostic and referral decisions for patients with vertigo. A lack of guidance contributes to the considerable variation in the investigation of patients with vertigo, with neuroimaging varying 8-fold between providers and admission rates ranging from 1-21% of patients.^10^ Currently, no individual or combination of clinical features accurately rules out a serious cause of vertigo or identifies which patients are at high risk for such a cause.^11^ This often leads to overuse of neuroimaging and prolonged emergency department stays for patients with benign dizziness, as well as missed or delayed diagnosis of serious conditions like stroke.^12–14^

Our study objectives were to prospectively assess the clinical characteristics of patients presenting with vertigo to the emergency department and to derive a clinical risk score to identify high and low-risk patients for a serious cause of their vertigo.

## Methods

### Study Design

This prospective multicenter cohort study was conducted in the emergency departments of three university-affiliated urban Canadian tertiary care teaching hospitals from July 2019 to August 2022.

### Study Population

We enrolled consecutive alert patients 18 years and older, who presented to participating emergency departments with a chief complaint of acute vertigo, dizziness or imbalance, and were assessed by an emergency department physician. Patients with symptom onset more than 14 days prior, head or neck trauma in the preceding 14 days, Glasgow Coma Scale score less than 15, systolic blood pressure less than 90mmHg, a syncopal episode in the preceding 14 days, or active cancer were excluded from the study. The research ethics board at each participating center approved the study without requiring written consent. Participants were informed that they might be contacted by telephone for follow-up, and verbal consent was obtained from such patients at telephone contact.

### Data Collection

Attending emergency physicians or supervised residents in emergency medicine completed all assessments. Physicians completed data forms to identify the presence or absence of 67 clinical findings in consecutive patients with vertigo. Variables included characteristics of the current event, physical examination findings, and medical history provided by the patient.

Research staff collected data forms, verified data, confirmed eligibility, and recorded objective data from physician, nursing, consultant, triage, ambulance, follow-up neurological consultations, and radiology reports. Objective data included – age, sex, date of visit, and documented ED diagnosis. Information was sought from the study hospital’s electronic medical records to identify subsequent emergency department visits, stroke/neurology clinic visits, and diagnostic imaging. For chart abstraction, a single trained reviewer at each stage abstracted data using a standardized data collection sheet. Chart abstractors underwent training(didactic session and 5 charts joint review) and testing(10 charts dual independent review); when testing resulted in a Kappa of >0.8 between the trainer and tester, they were validated for independent data abstraction. We conducted telephone follow-up calls at 7, 30 and 90 days to assess for subsequent stroke, TIAs, vertebral artery dissection, or brain tumour diagnoses. We used a previously validated tool, the Questionnaire for Verifying Stroke-Free Status to assess for outcomes.^15^ In addition, during the telephone follow-up call, patients were asked whether they were admitted to hospital at any point after their initial emergency department visit. If they were, they were asked for what condition and which symptoms they had, the duration of their symptoms, date of symptom onset, and which side was affected (if applicable).

Study staff reviewed emergency department census reports to identify any possible missed patients. If the eligibility criteria did not exclude patients, they were deemed potential missed patients. Data were entered into a computerized database using Statistical Analysis System (SAS) software. Data management and study coordination were conducted at the Health Sciences North Research Institute.

### Variables

We collected data on 67 different clinical variables. A priori, we identified clinically significant variables that were known to be associated with one or more of our outcomes; Age, Sex, hypertension, previous stroke, diabetes, atrial fibrillation, motor/sensory deficits, diplopia, dysarthria, dysphagia, dysmetria, ataxia and those that were likely to be negatively associated with a serious diagnosis ( BPPV, multiple episodes). These were candidate variables for the model. A scoping review and expert opinion informed the choice of these variables. Emergency department clinician diagnosis of a serious outcome: this was defined as the documented discharge or admission diagnosis of stroke, TIA, vertebral artery dissection or brain tumour.

Serious diagnosis on computed tomography: this was defined as radiological reported evidence of acute stroke (ischemic or hemorrhagic), vertebral artery dissection or brain tumour/mass.

### Outcome Measures

The primary outcome of a serious diagnosis was defined as a diagnosis of stroke, TIA, vertebral artery dissection, or brain tumour diagnosed in the emergency department or within 30 days of the initial assessment. Outcomes were defined as follows. Stroke (ischemic and hemorrhagic): rapidly developed clinical symptom(s) of focal (or occasionally global) disturbance of cerebral function lasting more than 24 hours or until death with no apparent non-vascular cause.^16^ TIA: sudden, focal neurological deficit lasting for less than 24 hours, presumed to be of vascular origin, and confined to an area of the brain or eye perfused by a specific artery.^16^ Brain tumour: radiological evidence of an intracranial mass that another more likely diagnosis cannot explain that required intervention (medical or surgical) within 30 days of diagnosis. Vertebral artery dissection: radiological evidence of vertebral artery dissection, hematoma or pseudoaneurysm.

#### Outcome Assessment

The primary outcome was assessed for all patients from a composite of sources, including site hospital records, autopsy reports at the site hospital, or patients who answered "yes" to at least one telephone follow-up question. An Adjudication Committee, blinded to the initial emergency department visit, reviewed all possible outcome events. The Adjudication Committee comprised three members: a stroke neurologist and two experienced emergency physicians. These assessors independently evaluated each possible outcome, and an event was considered to have occurred if at least two of the three physicians agreed. Secondary outcomes followed a similar process.

### Data Analysis

Descriptive statistics were computed using frequencies and proportions for categorical variables and means and standard deviations for continuous variables. We compared proportions and mean differences using the chi-square test or fisher exact test as appropriate and t-test, respectively. A two-sided p-value below 0.05 suggested a statistically significant difference.

Univariate and multivariate logistic regression analysis were used to assess the association between predictors and outcome. A multivariate logistic regression model was developed. Initial variable selection was based on unadjusted significant associations with the outcome (p<0.20) and clinical significance. Clinical significance was based on previous systematic reviews and expert opinion. The variables were evaluated for effect modification using interaction terms and collinearity using computed collinearity diagnostics (i.e. Condition index, variance inflation factor). Our final multivariate logistic regression model included the most important and significant variables. We assigned points to our final model predictors by dividing the beta coefficients of the predictors by the smallest of the beta coefficients and rounding the decimal quotients to the nearest integer. This was to simplify the calculation and increase usability. We calculated the total score for each patient.

Internal validation of the model was carried out with bootstrapping, in which we used 1000 bootstrap samples sampled randomly with replacement. The optimism and optimism-correct C-statistic were calculated. We assessed the calibration of the model and score using a calibration slope between observed and predicted probabilities at each score category. Because of the small number of patients and events in the higher risk scores, we collapsed the scores above 14.

We compared the score’s ability to discriminate between outcomes and non-outcomes to previously derived decision aids/scores (TriAGe +, STANDING algorithm, DEFENSIVE stroke scale, the nomogram for stroke risk assessment), clinical judgment and computed tomography. Discrimination was quantified with the concordance (*c*)-statistic. A significant difference between tools was assessed using the DeLong method, which compares two area under the curve (AUC) values.

Where more than one variable data was missing from a patient, they were excluded from the analysis.

We assessed the impact of the score on resource utilization using the score level that would define a low-risk group with 0 serious diagnoses. In order to provide the most conservative estimate, we assumed no CT would be performed in the low-risk group, but every patient in the medium and high-risk groups would now undergo a CT. This is unlikely how the risk score would be used, and therefore, this is an underestimation of the expected decrease in CT utilization.

All statistical analyses were performed using SAS software version 9.4.

Based on the method proposed by Riley et al., we estimated a required sample size of 85 outcome events.^17^ This was based on the assumption of a shrinkage of 0.9, Cox-Snell R squared of 0.1, an outcome proportion rate between 0.02 and 0.05 and a model based on up to 20 predictors.

## Results

In this study, we enrolled 2078 of 2618 potentially eligible patients (79.4%) There were 2 patients missing sex and age, they were excluded. We had 4 patients who were missing diastolic blood pressure (Figure 1). We had a mean age of 77.1 years, with 59% women. A CT head scan was performed in 643 (30.9%) patients and a magnetic resonance imaging (MRI) scan in 56 (2.5%) patients. There were 160 (7.7%) admitted to the hospital and specialist consultation was requested (in emergency or as outpatient) for 234 (11.3%) patients. There were 111 (5.3%) serious diagnoses, including stroke 99 (81.1%), TIA 11 (9.9%), vertebral artery dissection 2 (1.8%), and brain tumour 1 (0.9%). Follow-up was complete for 80.4% of the cohort at 30 days.

**Figure 1.**
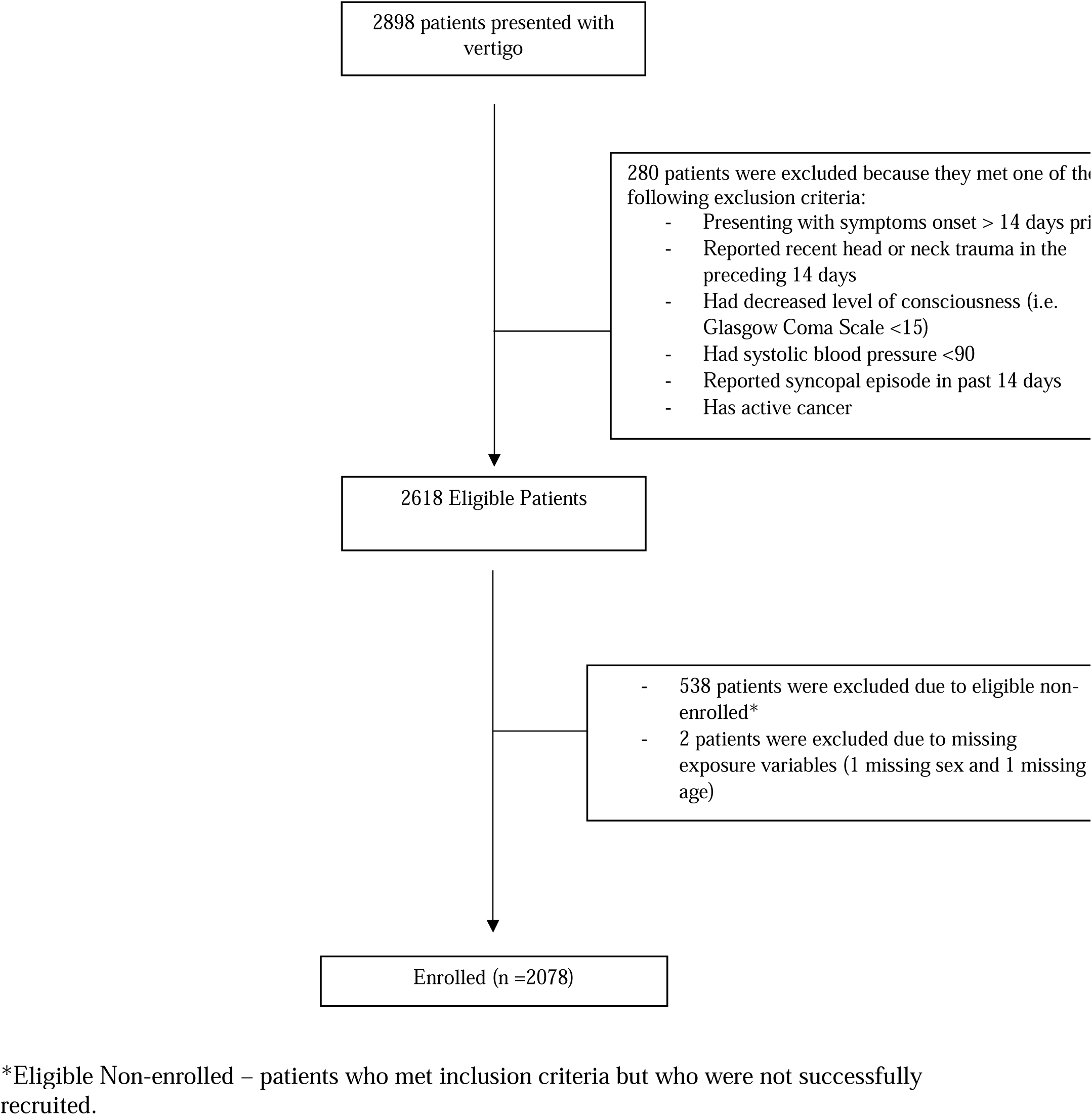
Flow Diagram of the Study Cohort

Table 1 reports the characteristics of enrolled patients. Clinical features strongly correlated with having a serious cause of vertigo included age greater than 65 years, mean systolic blood pressure (157 mmHg), previous stroke, previous TIA, hypertension, diabetes, dysphagia, motor deficit, sensory deficit, ataxia, dysarthria, ongoing dizziness, dizziness lasting more than 2 minutes or multiple episodes of dizziness. Dizziness triggered by a change in any position and a clinical diagnosis of benign paroxysmal positional vertigo were negatively associated with a serious diagnosis.

**Table 1.** Baseline Characteristics of Patients Presenting to Emergency Department with Dizziness According to Stroke Events.

| Predictors<sup>*</sup> |  | Central Outcome (n=2078) |  | P-value<sup>†</sup> |
| --- | --- | --- | --- | --- |
|  |  | Yes (n=111) | No (n=1967) |  |
| Sex (%), Female |  | 46 (41.4) | 1185 (60.2) | <0.0001 |
| Age 65 or Over |  | 84 (75.7) | 751 (38.2) | <0.0001 |
| Mean Heart Rate, bpm (SD)<sup>2</sup> |  | 79.89 (17.2) | 79.03 (15.2) | 0.5640 |
| Mean Systolic Blood Pressure, mmHg (SD) |  | 156.1 (28.5) | 138.30 (24.2) | <0.0001 |
| Mean Diastolic Blood Pressure, mmHg (SD) |  | 82.43 (14.7) | 81.31 (12.0) | 0.3532 |
| Past Medical History | Previous Stroke | 30 (27.0) | 89 (4.5) | <0.0001 |
|  | Previous Transient Ischemic Attack | 9 (8.1) | 51 (2.6) | 0.0038 |
|  | Hypertension | 100 (90.1) | 1214 (61.7) | <0.0001 |
|  | Diabetes | 34 (30.6) | 254 (12.9) | <0.0001 |
|  | Atrial Fibrillation | 10 (9.0) | 113 (5.7) | 0.1562 |
| Neurological Deficits | Dysphagia | 6 (5.4) | 9 (0.5) | <0.0001 |
|  | Diplopia | 14 (12.6) | 48 (2.4) | <0.0001 |
|  | Motor Deficit | 45 (40.5) | 28 (1.4) | <0.0001 |
|  | Sensory Deficit | 17 (15.3) | 23 (1.2) | <0.0001 |
|  | Ataxia | 61 (55.0) | 151 (7.7) | <0.0001 |
|  | Dysarthria | 34 (30.6) | 12 (0.6) | <0.0001 |
|  | Dysmetria | 28 (25.2) | 27 (1.4) | <0.0001 |
| Symptoms | Nausea | 46 (41.4) | 941 (47.8) | 0.1891 |
|  | Vomiting | 28 (25.2) | 410 (20.8) | 0.2708 |
|  | Headache | 40 (36.0) | 536 (27.3) | 0.0442 |
|  | Neck Pain or Discomfort | 3 (2.7) | 126 (6.4) | 0.1157 |
|  | Facial Eye Pain | 4 (3.6) | 46 (2.3) | 0.3385 |
|  | Hearing Loss | 0 | 48 (2.4) | 0.1093 |
|  | Tinnitus | 3 (2.7) | 140 (7.1) | 0.0738 |
|  | Recent Viral Upper Respiratory Tract Infection Symptoms | 4 (3.6) | 111 (5.6) | 0.3605 |
|  | Unable to Walk Unaided | 42 (37.8) | 71 (3.6) | <0.0001 |
|  | Can Walk More than 10 Steps | 17 (15.3) | 434 (22.1) | 0.0933 |
|  | Nystagmus | 14 (12.6) | 177 (9.0) | 0.1997 |
| Timing | Ongoing | 69 (62.2) | 625 (31.8) | <0.0001 |
|  | Gradual | 12 (10.8) | 347 (17.6) | 0.0640 |
|  | Abrupt | 89 (80.2) | 1484 (75.4) | 0.2578 |
|  | More than 2 mins | 88 (79.3) | 1033 (52.5) | <0.0001 |
| Episodes | Single | 82 (73.9) | 757 (38.5) | <0.0001 |
|  | Multiple | 23 (20.7) | 1173 (59.6) | <0.0001 |
| Movement Triggers | Head Turning | 6 (5.4) | 437 (22.2) | <0.0001 |
|  | Getting Up | 18 (16.2) | 523 (26.6) | 0.0154 |
|  | Lying Down | 1 (0.9) | 157 (8.0) | 0.0062 |
|  | Bending Over | 2 (1.8) | 118 (6.0) | 0.0651 |
|  | Looking Up | 1 (0.9) | 51 (2.6) | 0.5236 |
|  | Rolling Over in Bed | 1 (0.9) | 150 (7.6) | 0.0079 |
|  | Walking | 10 (9.0) | 138 (7.0) | 0.4270 |
|  | Any | 6 (5.4) | 278 (14.1) | 0.0092 |
|  | Persistent When Still | 6 (5.4) | 151 (7.7) | 0.3784 |

<sup>&</sup>frequency of missing in event group: 4 diastolic blood pressure
\*comparing the presence of predictor to absence (using “no” as reference category)
<sup>1</sup>p-values determined using two-sample t-test for continuous predictors, chi-square test (for categorical predictors with expected cell sizes over 5) and the fisher exact test (for categorical predictors with expected cell sizes below 5)
<sup>2</sup> missing observations for heart rate (n=15), systolic blood pressure (n=11) and diastolic blood pressure (n=87)

Our multivariate analysis calculated adjusted odds ratios for clinical variables deemed to be clinically significant (Table A-1). We found six variables independently positively associated with a serious diagnosis. Benign paroxysmal positional vertigo was negatively associated with a serious diagnosis. The model had excellent discrimination (C-statistic of 0.972) (Figure 2), which remained unchanged when we adjusted for optimism (C-statistic of 0.969).

**Figure 2.**
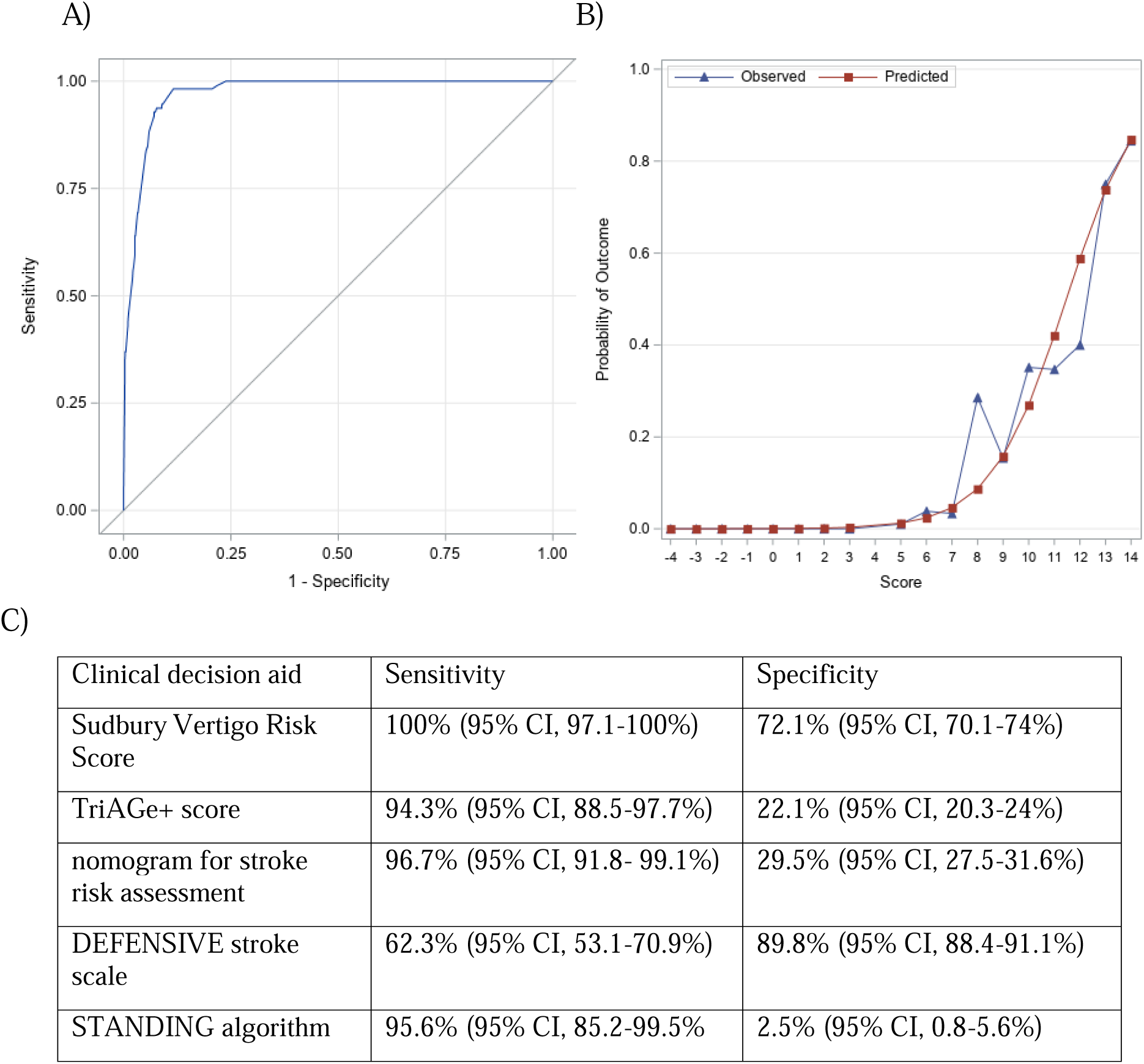
A) ROC Curve of Multivariate Logistic Regression Model B) Observed and Predicted Probabilities of a Serious Diagnosis by Score C) Diagnostic accuracy of the Sudbury Vertigo Risk Score compared to alternative decision aids.

Table 2 lists the seven components of the Sudbury Vertigo Risk Score obtained from the clinical history and examination. The total score ranges from −4 to 17. The probability of a serious cause ranged from 0% for a score of <5, 2.1% for a score of 5-8, and 41% for a score >8 (Table 3). Our score showed good calibration between the observed and predicted probabilities of a serious diagnosis at each score category (Figure 2). For our primary outcome, a serious diagnosis, the sensitivity was 100% (95% CI, 97-100%) and the specificity 72.1% (95% CI, 70.1-74%) for a score >4. Using a score of >4 to define a high-risk group that warrants further investigation would reduce CT use by 10%.

**Table 2.**
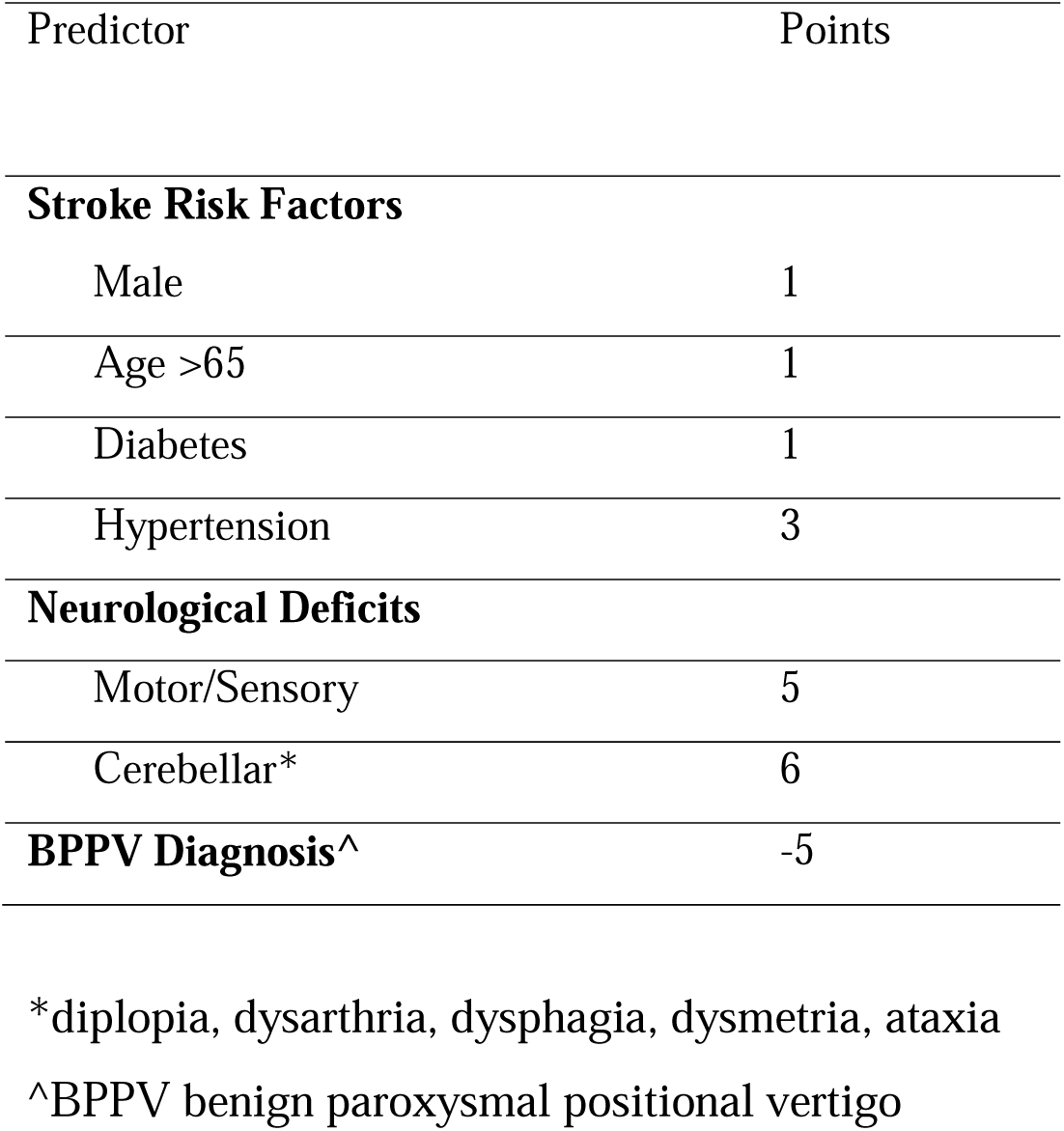
Sudbury Vertigo Risk Score.

**Table 3.**
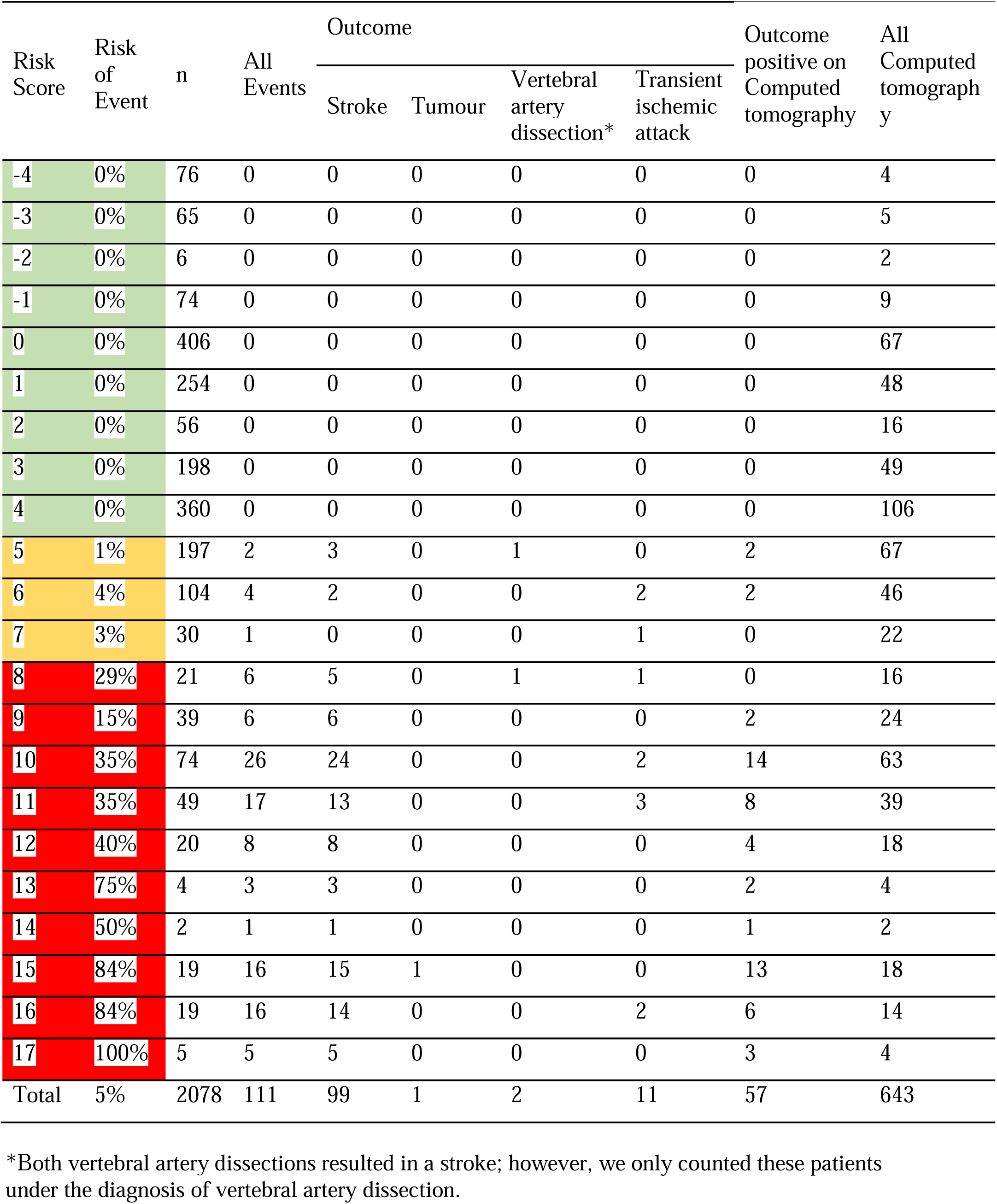
Patients, serious outcomes and image findings at each score level of the Sudbury Vertigo Risk Score.

| Risk Score | Risk of Event | n | All Events | Outcome |  |  |  | Outcome positive on Computed tomography | All Computed tomography |
| --- | --- | --- | --- | --- | --- | --- | --- | --- | --- |
|  |  |  |  | Stroke | Tumour | Vertebral artery dissection* | Transient ischemic attack |  |  |
| -4 | 0% | 76 | 0 | 0 | 0 | 0 | 0 | 0 | 4 |
| -3 | 0% | 65 | 0 | 0 | 0 | 0 | 0 | 0 | 5 |
| -2 | 0% | 6 | 0 | 0 | 0 | 0 | 0 | 0 | 2 |
| -1 | 0% | 74 | 0 | 0 | 0 | 0 | 0 | 0 | 9 |
| 0 | 0% | 406 | 0 | 0 | 0 | 0 | 0 | 0 | 67 |
| 1 | 0% | 254 | 0 | 0 | 0 | 0 | 0 | 0 | 48 |
| 2 | 0% | 56 | 0 | 0 | 0 | 0 | 0 | 0 | 16 |
| 3 | 0% | 198 | 0 | 0 | 0 | 0 | 0 | 0 | 49 |
| 4 | 0% | 360 | 0 | 0 | 0 | 0 | 0 | 0 | 106 |
| 5 | 1% | 197 | 2 | 3 | 0 | 1 | 0 | 2 | 67 |
| 6 | 4% | 104 | 4 | 2 | 0 | 0 | 2 | 2 | 46 |
| 7 | 3% | 30 | 1 | 0 | 0 | 0 | 1 | 0 | 22 |
| 8 | 29% | 21 | 6 | 5 | 0 | 1 | 1 | 0 | 16 |
| 9 | 15% | 39 | 6 | 6 | 0 | 0 | 0 | 2 | 24 |
| 10 | 35% | 74 | 26 | 24 | 0 | 0 | 2 | 14 | 63 |
| 11 | 35% | 49 | 17 | 13 | 0 | 0 | 3 | 8 | 39 |
| 12 | 40% | 20 | 8 | 8 | 0 | 0 | 0 | 4 | 18 |
| 13 | 75% | 4 | 3 | 3 | 0 | 0 | 0 | 2 | 4 |
| 14 | 50% | 2 | 1 | 1 | 0 | 0 | 0 | 1 | 2 |
| 15 | 84% | 19 | 16 | 15 | 1 | 0 | 0 | 13 | 18 |
| 16 | 84% | 19 | 16 | 14 | 0 | 0 | 2 | 6 | 14 |
| 17 | 100% | 5 | 5 | 5 | 0 | 0 | 0 | 3 | 4 |
| Total | 5% | 2078 | 111 | 99 | 1 | 2 | 11 | 57 | 643 |

\*Both vertebral artery dissections resulted in a stroke; however, we only counted these patients under the diagnosis of vertebral artery dissection.

CT had a sensitivity of 45.9% (95% CI 36.8 -55.2%) and a specificity 100% (95% CI, 99-100%) for a serious outcome. Clinical judgment had a sensitivity of 82% (95% CI, 73.6-88.6%) and a specificity of 99.4% (95% CI, 99-99.7%).

Using the DeLong method, we found that the derived clinical risk score had significantly better discrimination (p<0.001) than previously published tools (TriAGe+ score, the nomogram for stroke risk assessment, DEFENSIVE stroke scale, and STANDING algorithm (Figure 2).

## Discussion

We derived a clinical risk score that can identify the risk for a serious diagnosis in a patient presenting with vertigo. The Sudbury Vertigo Risk Score can be used to aid in identifying the subset of patients at low risk who can be safely discharged without further investigation, referral, or admission and triage those at high risk for urgent testing and treatment. The risk score could potentially reduce unnecessary healthcare costs and prevent missed or delayed diagnosis of serious diagnoses.

## Previous Studies

There are no clinical risk scores or decision aids with sufficient sensitivity to rule out a serious diagnosis in vertigo patients. In two surveys, emergency department physicians reported needing a clinical risk score to help assess vertigo patients. They defined a required miss rate of <1%.^18,19^ Six clinical decision-aids/scores have been derived, all subject to small sample sizes, a high risk of bias or unacceptable accuracy.^20–24^ The *HINTS exam* incorporates three physician exam assessments: the head impulse test, nystagmus and test of skew. However, it only applies to those presenting with acute vestibular syndrome (a subset of patients with constant vertigo, head motion intolerance, nystagmus, ataxia, and nausea/vomiting). Acute vestibular syndrome accounts for only 10% of those presenting with vertigo.^25,26^ It has failed validation for use by emergency physicians, with sensitivity ranging from 66.7-85%.^25,26^ The *STANDING algorithm* consists of the (1) discrimination between spontaneous and positional nystagmus, (2) evaluation of the nystagmus direction, (3) head impulse test, and (4) evaluation of equilibrium, the second and third of which are components of the HINTS exam.^21^ On external validation, the sensitivity was only 93.6%, which was similar in our cohort (95.5%) but with a significantly lower specificity of 2.5%. This is likely related to the difficulty in performing its components, with all physicians rating confidence in assessing nystagmus and head impulse test as low.^18,27^ The *TriAGe+ score* consists of eight variables: triggers, atrial fibrillation, male, hypertension, brainstem or cerebellar dysfunction, focal weakness or speech impairment, dizziness and no history of vertigo. In our cohort, we found a sensitivity of 95.3% and a specificity of 22.1%.^22^ The *nomogram for stroke-risk assessment* is based on sex, trigger, isolated symptoms, nausea, history of brief dizziness, high blood pressure, finger-nose test and tandem gait assessment. In the derivation study with components assessed by a neurologist, it demonstrated a high diagnostic accuracy. In our cohort, the sensitivity was 96.7% and the specificity 29.5%. Other clinical decision aids derived based on neurology-assessed clinical variables have failed prospective validation, with significantly lower diagnostic accuracy when performed by emergency department physicians.^25,26,28^ Yamada et al., through assessment of a case series of posterior circulation strokes, decided upon a three-item checklist called the *DEFENSIVE stroke scale*; sensory disturbance, ataxia or visual deficit. Internal validation found a sensitivity of 100%. This retrospective study suffered from spectrum bias with a high percentage of serious outcomes(9.7%) in the cohort, and had incomplete patient outcome assessment. In our cohort, we found a sensitivity 62.3% and a specificity of 89.8%. Our cohort had a 5% prevalnce of serious diagnosis, this is more represenative of other emergency departmetn studies on vertigo patients.^5,29,30^ All tools had a lower area under the curve than the newly derived score.

## Clinical Implications

The Sudbury Vertigo Risk Score can likely be categorized into strata that dictate a course of action. This may include no further investigation for low risk patients (e.g., <1% risk of a serious diagnosis, score <5), further investigation for moderate risk if no alternative diagnosis (1-5%, score 5-8) and expedited/ same-visit consultation and investigation for those at high risk (e.g., >5%, score >8).

Patients with a score of <5 are low-risk based on this study; if a clinician’s judgement is concurrent with this, they can be further reassured that their patient is low-risk. Patients with low-risk scores for which clinicians have a moderate to high pretest probability for a serious etiology should follow their clinical judgement until the score is further validated.

Patients deemed to be high-risk with a score of >8 should be considered high-risk and managed as such. This group should be prioritized for urgent testing and treatment, 80% of missed cases were within this group. These missed patients represent a population whose care can be improved now by this risk score without increasing the investigation of low-risk patients.

Further investigation should be based on clinical judgment for those deemed moderate risk. If there is no alternative diagnosis, a serious diagnosis should be considered, and further investigation/referral/treatment should be initiated.

Over a third of patients underwent CT. Using CT as a test to rule out a serious diagnosis is of limited benefit with a low sensitivity.^8^ We found a sensitivity of 45.9% (95% CI 36.8 -55.2%) in our cohort. This is higher than a recent systematic review by Shah et al. They found a pooled sensitivity of 28*.5% (95% CI 14.4%–48.5%)* and a of specificity 98.9*% (95% CI 93.4%– 99.8%,).* ^8,31,32^ Not performing CT in those with a score <5 would reduce CT usage by >50%. However, this would be an overly optimistic estimate as the use of any such score would likely increase CT usage in those deemed to be at risk for a serious diagnosis. Even if all those with a score ≥5 were to undergo a CT this would still decrease CT use by 10% and still identify all positive findings.

## Strengths

We conducted a large prospective multicenter cohort study of patients with vertigo. Our study enrolled a representative sample of patients presenting with vertigo, addressing the spectrum bias seen in previous decision tool derivation studies. This study prospectively assessed history and examination findings to identify patients at high risk for a serious diagnosis. We also followed the methodological standards recommended for derivation studies for clinical decision rules.^33,34^ These standards allow for a more reproducible way of deriving a risk score, resulting in a more robust tool than consensus-based risk scores. Our score used variables available to clinicians at the bedside. Our study primarily enrolled patients diagnosed by frontline emergency physicians, which allows our results to be highly generalizable. All physicians enrolling patients were well-trained, certified emergency medicine specialists, reducing classification bias. Our use of blinded Adjudication Committees to assess subsequent serious diagnoses provided a highly rigorous event classification.

## Limitations

We used the World Health Organisation (WHO) definitions of stroke and TIA. These are clinical diagnoses mainly based on history and examination without the benefit of MRI to exclude small infarcts. This is consistent with current practice in most emergency departments.^38^ However, it may overestimate the actual number of ischemic strokes by including stroke mimics.^35^ These patients have stroke-like symptoms due to other etiologies. Given the lack of immediate MRI in our study centres for all these patients, we utilized the WHO definition.

Not all eligible patients were enrolled. We do not suspect any systematic reason for this other than the realities of conducting research in busy, tertiary care emergency departments. Our results may not be generalized to emergency departments in different settings (i.e., rural, non-academic, community). We did not have complete follow-up data for 19.6% of our cohort. None of these had neurological deficits. Previous studies have identified the risk of a subsequent stroke in a population with isolated vertigo of <1%. However, we could have misclassified a patient with a serious diagnosis as a non-outcome. This could artificially increase the reported sensitivity.

## Conclusions

The Sudbury Vertigo Risk Score identifies the risk of a serious diagnosis as a cause of a patient’s vertigo. If validated, it could help guide physician investigation, consultation and treatment decisions, improving resource utilization and reducing missed diagnoses.

## Data Availability

Available on request

## Appendix

**Table A-1.** Univariate and Multivariate Logistic Regression Analysis of Central Cause Predictors in Patients Presenting to the Emergency Department with Dizziness (n=2028)

| Predictors |  | Crude OR | P-value | Adjusted OR | P-value |
| --- | --- | --- | --- | --- | --- |
| Sex (%), (ref=female) |  | 2.1 (1.5-3.2) | 0.0001 | 2.3 (1.3-4.2) | 0.0073* |
| Age | Below 65 | Reference |  | Reference |  |
|  | 65 or Over | 5.0 (3.2-7.9) | <.0001* | 2.7 (1.4-5.2) | 0.0043* |
| Past Medical History<sup>1</sup> | Previous Stroke | 7.8 (4.9-12.5) | <.0001* | -- | -- |
|  | Previous Transient Ischemic Attack | 3.3 (1.6-6.9) | 0.0014* | -- | -- |
|  | Hypertension | 5.6 (3.0-10.6) | <.0001* | 9.3 (3.1-27.8) | <.0001* |
|  | Diabetes | 3.0 (2.0-4.6) | <.0001* | 2.6 (1.3-4.9) | 0.0050* |
|  | Atrial Fibrillation | 1.6 (0.8-3.2) | 0.1601 | -- | -- |
| Neurological Deficits<sup>1</sup> | Dysphagia | 12.4 (4.3-35.6) | <.0001* | 16.2 (2.8-93.4) | 0.0018* |
|  | Diplopia | 5.8 (3.1-10.8) | <.0001* | 3.8 (1.5-9.3) | 0.0040* |
|  | Motor Deficit | 47.2 (27.7-80.4) | <.0001* | 16.8 (7.3-38.6) | <.0001* |
|  | Sensory Deficit | 15.3 (7.9-29.6) | <.0001* | 9.1 (3.0-27.8) | <.0001* |
|  | Ataxia | 14.7 (9.8-22.1) | <.0001* | 8.5 (4.5-16.0) | <.0001* |
|  | Dysarthria | 71.9 (35.9-144.3) | <.0001* | 94.1 (34.8-254.3) | <.0001* |
|  | Dysmetria | 24.2 (13.7-43.0) | <.0001* | 6.8 (2.9-16.3) | <.0001* |
| Symptoms<sup>2</sup> | Nausea | 0.8 (0.5-1.1) | 0.1902 | -- | -- |
|  | Vomiting | 1.3 (0.8-2.0) | 0.2717 | -- | -- |
|  | Headache | 1.5 (1.0-2.2) | 0.0455 | -- | -- |
|  | Neck Pain or Discomfort | 0.4 (0.1-1.3) | 0.1281 | -- | -- |
|  | Facial Eye Pain | 1.6 (0.6-4.4) | 0.4013 | -- | -- |
|  | Hearing Loss<sup>3</sup> | 0.3 (0.00-1.3) | 0.0884 | -- | -- |
|  | Tinnitus | 0.4 (0.1-1.2) | 0.0866 | -- | -- |
|  | Recent Viral URTI Symptoms | 0.6 (0.2-1.7) | 0.3649 | -- | -- |
|  | Unable to Walk Unaided | 16.3 (10.4-25.5) | <.0001* | -- | -- |
|  | Can Walk More than 10 Steps | 0.6 (0.4-1.1) | 0.0959 | -- | -- |
|  | Nystagmus | 1.5 (0.8-2.6) | 0.2022 | -- | -- |
| Timing | Ongoing | 3.5 (2.4-5.2) | <.0001* | -- | -- |
|  | Gradual | 0.6 (0.3-1.0) | 0.0674 | -- | -- |
|  | Abrupt | 1.3 (0.8-2.1) | 0.2594 | -- | -- |
|  | More than 2 mins | 3.5 (2.2-5.5) | <.0001* | -- | -- |
| Episodes | Single | 4.5 (2.9-7.0) | <.0001* | -- | -- |
|  | Multiple | 0.2 (0.1-0.3) | <.0001* | -- | -- |
| Movement Triggers | Head Turning | 0.2 (0.1-0.5) | <.0001* | -- | -- |
|  | Getting Up | 0.5 (0.3-0.9) | 0.0170* | -- | -- |
|  | Lying Down | 0.1 (0.02-0.8) | 0.0252* | -- | -- |
|  | Bending Over | 0.3 (0.1-1.2) | 0.0833 | -- | -- |
|  | Looking Up | 0.3 (0.1-2.5) | 0.2900 | -- | -- |
|  | Rolling Over in Bed | 0.1 (0.02-0.8) | 0.0286* | -- | -- |
|  | Walking | 1.3 (0.7-2.6) | 0.4276 | -- | -- |
|  | Any | 0.4 (0.2-0.8) | 0.0127* | -- | -- |
|  | Persistent When Still | 0.7 (0.3-1.6) | 0.3811 | -- | -- |
| BPPV Diagnosis |  | 0.03 (0.01-0.2) | 0.0005* | 0.1 (0.01-0.5) | 0.0093* |

\*significant association (alpha<0.05 for univariate and top 10 for multivariate); <sup>1</sup>reference categories are defined as no; hypertension defined as a history of or prevalent hypertension at presentation

